# Effect of Tranexamic acid for acute spontaneous intracerebral haemorrhage: A systematic review and individual patient data meta-analysis

**DOI:** 10.1101/2025.11.10.25339961

**Authors:** Chaamanti S. Menon, Zhe Kang Law, Lisa Woodhouse, Jingyi Liu, Chloe Mutimer, Michael Desborough, Liping Liu, Alexandros Polymeris, Atte Meretoja, Nawaf Yassi, Henry Zhao, Stephen Davis, Geoffrey Donnan, Rob A. Dineen, Jeyaraj Pandian, David Seiffge, Rustam Al-Shahi Salman, Philip M. Bath, Nikola Sprigg

## Abstract

**Background:** Spontaneous intracerebral haemorrhage (ICH) has high rates of death and disability, and no proven haemostatic treatment. Tranexamic acid might limit haematoma expansion and improve outcomes. We conducted the first individual patient data meta-analysis to evaluate the effect of tranexamic acid on functional outcomes in spontaneous ICH.

**Methods:** This systematic review and individual patient data meta-analysis included randomised controlled trials comparing intravenous tranexamic acid to placebo in adults with spontaneous ICH treated within 12 hours of onset. MEDLINE, EMBASE, CENTRAL, Web of Science, and WHO ICTRP were searched to November 2024. The primary outcome was functional status at 90 days after randomisation, measured by the modified Rankin Scale. Adverse events (seizures, thromboembolic events) were also summarised. Analyses used generalised linear mixed models with random intercepts for trial. Risk of bias was assessed using the Cochrane RoB 2 tool. The study was registered with PROSPERO (CRD42022345775).

**Results:** We identified 1,131 records; nine trials (3,194 participants) were eligible for inclusion and five trials (2860 participants; 90% of those available) provided individual patient data for the primary analysis. Risk of bias was low across all included trials. At 90 days, 757/1423 (53.2%) patients assigned to tranexamic acid had a worse functional outcome compared with 759/1415 (53.6%) assigned to placebo, the difference was not statistically significant (adjusted common odds ratio 0.93 (95% CI 0.81 to 1.07; p = 0.30). Serious adverse events were similar between the groups, with no significant differences observed. There was no evidence of between-trial heterogeneity based on model fit (likelihood ratio test).

**Discussion:** Completed clinical trials provide no evidence that tranexamic acid improves functional outcome after spontaneous ICH. However, given the reduction in hematoma expansion and early mortality it remains to be seen if this translates to improved functional outcome in larger ongoing clinical trials. Even a small beneficial effect could have potential for global impact given the burden of ICH.

**Funding:** No funding source.

## Introduction

Intracerebral haemorrhage (ICH) represents ∼20% of all strokes and is a leading cause of death and long-term disability^1^. Unlike ischaemic stroke, effective acute therapies for ICH remain limited, and outcomes are primarily dictated by the extent of haematoma expansion, a key driver of secondary brain injury. Haematoma expansion occurs in up to 40% of patients within the first 24 hours and is strongly associated with increased mortality and worse functional outcomes^2^.

The underlying mechanism of haematoma expansion is multifactorial, involving a process of ongoing bleeding from ruptured cerebral vessels. The initial insult triggers local coagulopathy, clot instability, and secondary fibrinolysis. Fibrinolysis is mediated by the conversion of plasminogen to plasmin, which degrades fibrin clots and exacerbates bleeding^3^. Additionally, the inflammatory response to haemorrhage further contributes to vascular permeability and clot breakdown, extending the cycle of expansion^4^.

Tranexamic acid, a synthetic lysine analogue, inhibits plasminogen activation and plasmin binding to fibrin, thereby stabilising the clot and reducing ongoing bleeding. This antifibrinolytic action has been demonstrated across various conditions, including trauma CRASH-2 trial)^5,6^ and postpartum haemorrhage (WOMAN trial)^7^. In the context of ICH, tranexamic acid holds promise in halting early haematoma expansion, which is critical in the hyperacute phase. Evidence from trials such as TICH-2 suggests tranexamic acid may reduce haematoma growth, although its impact on clinical outcomes remains uncertain.

This study is the first systematic review and individual patient data meta-analysis to pool data across multiple trials to assess the efficacy of tranexamic acid in spontaneous intracerebral haemorrhage, focusing on functional independence, mortality, and haematoma expansion

## Methods

This systematic review with individual patient data meta-analysis was prospectively registered and the protocol has been published on PROSPERO (CRD42022345775)^8^. The review is reported in accordance with the PRISMA-IPD statement. The initial protocol was published in 2017, with an update amendment and database lock in 2025 prior to the final analysis^8^. A comprehensive search was conducted to identify randomised trials of tranexamic acid in spontaneous ICH. The individual patient data meta-analysis was designed to assess the effects of tranexamic acid on functional and radiological outcomes in patients with spontaneous ICH.

### Search Strategy and Selection Criteria

Searches were conducted in MEDLINE, EMBASE, Cochrane CENTRAL, and Web of Science in November 2024. The search terms included “intracerebral haemorrhage,” “tranexamic acid,” “antifibrinolytic agents,” and “haemostatic agents.” Additional searches were carried out in ClinicalTrials.gov and the WHO International Clinical Trials Registry Platform (WHO ICTRP). The reference lists of relevant trials were also screened. Full details of the search strategy are provided in the supplementary data.

All titles and abstracts were screened by one reviewer to identify eligible studies. Full texts were retrieved for potentially relevant articles and assessed for inclusion. Trial eligibility was confirmed by reviewing published protocols, contacting trial investigators, and examining data sharing availability. Where individual participant data was not available, aggregated outcome data were included in a secondary analysis. Eligible trials were randomised controlled trials comparing tranexamic acid with placebo in patients with acute spontaneous ICH, with treatment initiated within 12 hours of symptom onset. Trials were excluded if they focused on traumatic or secondary causes of ICH, such as arteriovenous malformations, tumours, or aneurysms, except for those related to anticoagulation.

### Data Collection and Risk of Bias Assessment

Principal investigators of eligible trials were contacted and invited to share anonymised individual patient data, trial protocols, and related publications. Data were accepted in various formats and were checked for completeness and accuracy against published results. Any discrepancies, outliers, or missing data were queried with the original trialists. Variables were harmonised to enable consistent analysis across trials. Continuous variables were retained in their original form where possible, and categorical variables were recoded using standard definitions. Haematoma growth was defined as a relative increase in haematoma volume of more than 33% or an absolute increase greater than 6 mL on imaging performed within 24 ± 12 hours after randomisation. Early neurological deterioration was defined as a drop of two or more points in the Glasgow Coma Scale (GCS) or an increase of four or more points in the National Institutes of Health Stroke Scale (NIHSS). Haematoma location was categorised as supratentorial deep, supratentorial lobar, or infratentorial.

Risk of bias for each included trial was assessed independently by two reviewers (CSM and JL) using the Cochrane Risk of Bias 2 tool^9^. Disagreements were resolved by consensus or by discussion with a third reviewer (MD). The quality of the evidence was further evaluated using the GRADE framework (CSM and JL).

### Outcomes

The primary outcome was functional status at 90 days, measured using the modified Rankin Scale. Secondary outcomes included mortality at day 7, 28, and 90, haematoma growth, thromboembolic events, seizures, and the identification of patient subgroups that may benefit from tranexamic acid. Pre-specified effect modifiers included age, sex, haematoma size and location, intraventricular haemorrhage, baseline systolic blood pressure, onset to treatment, and prior use of antithrombotic therapy^8^.

### Statistical Analysis

Baseline characteristics were summarised as counts and percentages for categorical variables, and as mean (standard deviation) or median (interquartile range) for continuous variables. No statistical comparisons between groups were performed, as all included studies were randomised trials; any observed differences were assumed to be due to chance.

A one-stage individual patient data meta-analysis was performed using generalised linear mixed models in SPSS (version 29). The models included a random intercept for trial to account for clustering and between-trial heterogeneity. Heterogeneity was assessed by estimating the variance of the random intercept and formally tested using a likelihood ratio test. The difference in −2 log likelihoods was compared against a chi-squared distribution with one degree of freedom. Binary outcomes were analysed using a binomial distribution with a log link function to estimate risk ratios and 95% confidence intervals. Ordinal outcomes were analysed using cumulative logit (proportional odds) models assuming a multinomial distribution, and results were presented as common odds ratios with 95% confidence intervals. The proportional odds assumption was assessed using a likelihood ratio test comparing the proportional odds model to a non-proportional alternative. Haematoma volume absolute difference was analysed using Quade non-parametric ANCOVA. Survival analysis was performed using Cox regression survival curves based on the date of death, where available, to compare survival between treatment and placebo arms. Subgroup analyses tested for interactions and stratified effects. All subgroup models included the same adjustment variables and retained the random intercept for trial. The forest plot was created with R software, version 4.2.2, using the ‘forestplot’ package. Missing data was not imputed.

As a prespecified secondary analysis^8^, a two-step meta-analysis was performed to include trials that were unable to share individual patient data ^10,11^. For these trials, treatment effect estimates were extracted from publications. These were combined with summary results from individual patient data trials using a random-effects model in RevMan. Between-study heterogeneity was assessed using the I² statistic.

Sensitivity analyses were conducted to test the robustness of findings. These included analysis of the 90-day modified Rankin Score as a binary outcome (0–3 vs 4–6) and included early deaths (within 24 hours) as a surrogate for haematoma expansion. The pre-specified random-effects model was retained for the primary analysis, with a fixed-effect model analysis done to confirm robustness of the findings given the evidence of no heterogeneity.

### Ethical Considerations

Ethical approval was not required as only anonymised data from previously published trials were used. Data sharing agreements were established with all collaborating trialists, and all data were securely stored in line with data protection regulations.

## Results

### Study Identification and Selection

A total of 1,131 records were identified through database searches, including EMBASE (n=447), MEDLINE (n=462), CENTRAL (n=137), Web of Science (n=72), and ICTRP (n=13). Three additional trials were identified from other systematic reviews. After removing 273 duplicates and excluding 844 irrelevant records based on title and abstract, 17 full-text trials were screened for eligibility. Of these, nine trials met criteria for individual participant data requests, totalling 3194 participants. individual patient data were successfully obtained from five randomised controlled trials, contributing a combined total of 2,860 participants (94% of all participants). The remaining four trials did not provide individual patient data due to lack of author response, ongoing publication, or language barriers. Relevant aggregate data were available for seven, comprising 3,040 participants in total. One trial was excluded because participants were recruited beyond 12 hours from symptom onset, and seven trials were ongoing at the time of screening (Figure 1).

**Figure 1.**
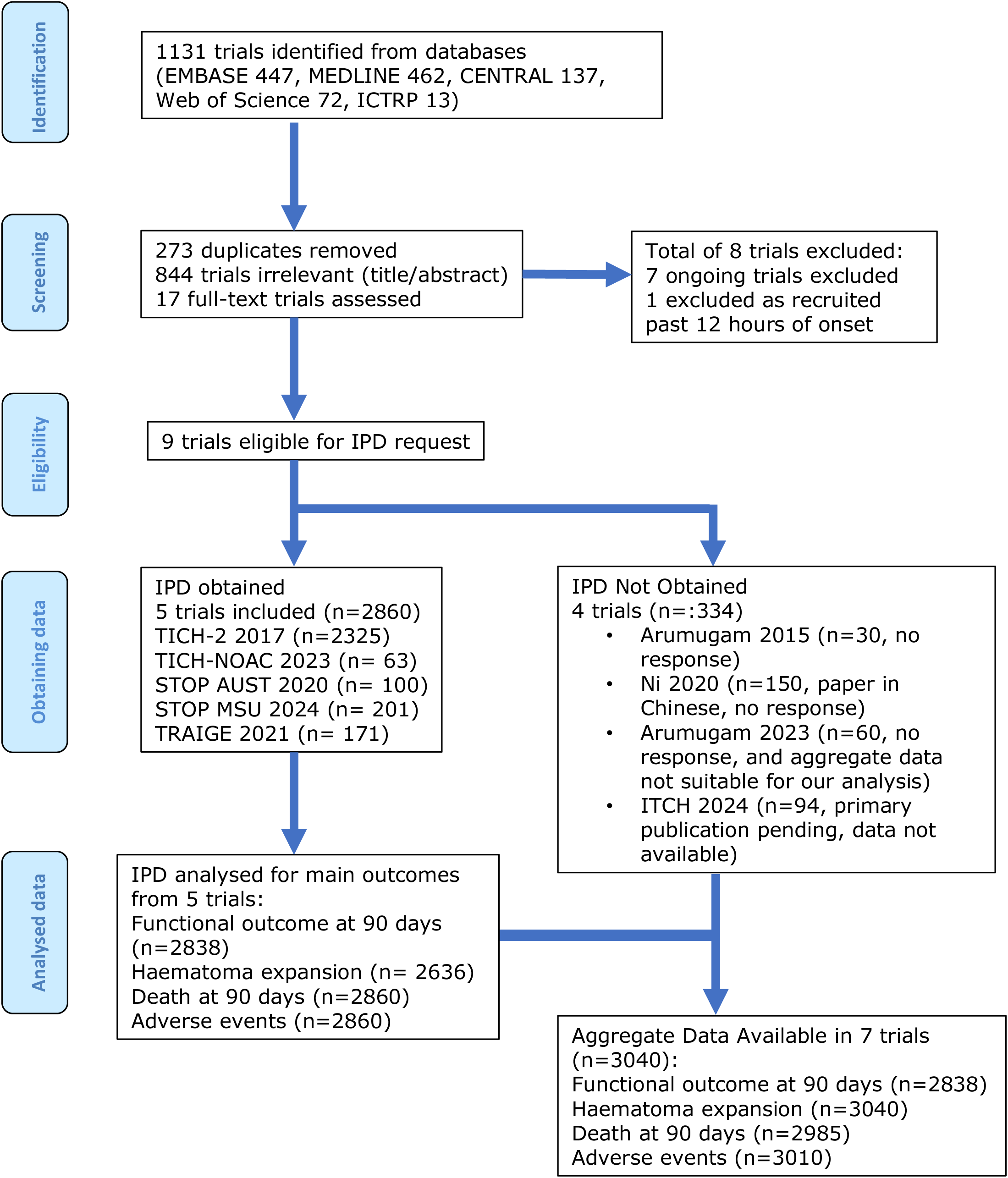
Study selection consort diagram (PRISMA-IPD adapted) Flow diagram showing the identification, screening, eligibility, and inclusion of trials for the individual participant data meta-analysis. Reasons for exclusion are indicated.

### Study Characteristics

The five included trials were conducted in multiple countries between 2017 and 2024. All trials investigated the effect of 2 g intravenous tranexamic acid administered within 2 to 12 hours of ICH onset. Most trials also included baseline CT imaging and follow-up CT at 24 hours. The trials also incorporated baseline CT angiography. All included trials were judged to be at low risk of bias and used blinded outcome assessment (Supplementary Figure 1). A summary of trial characteristics is provided in Table 1^12–16^. There were no important issues identified when checking individual patient data.

**Table 1.**
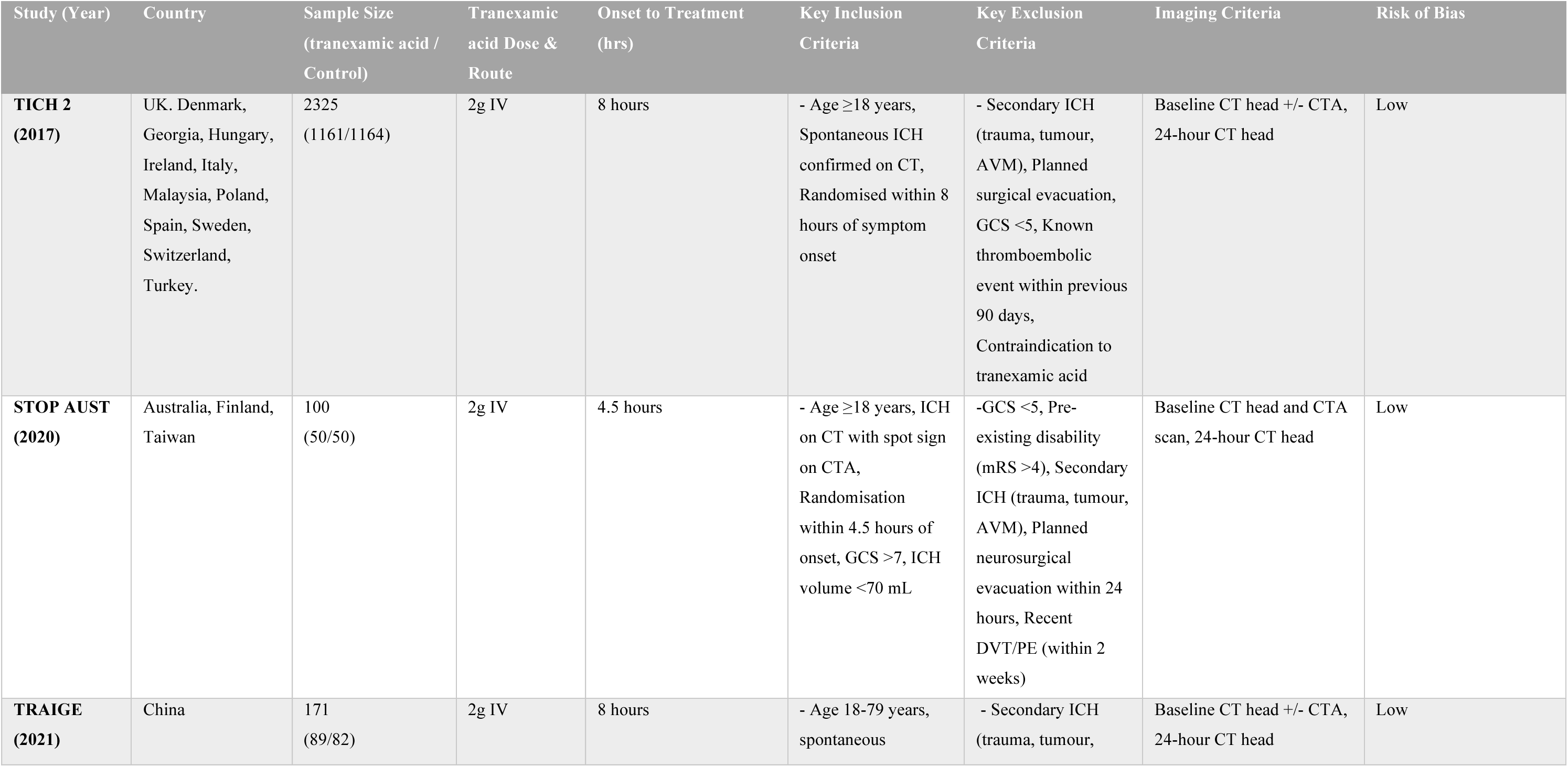
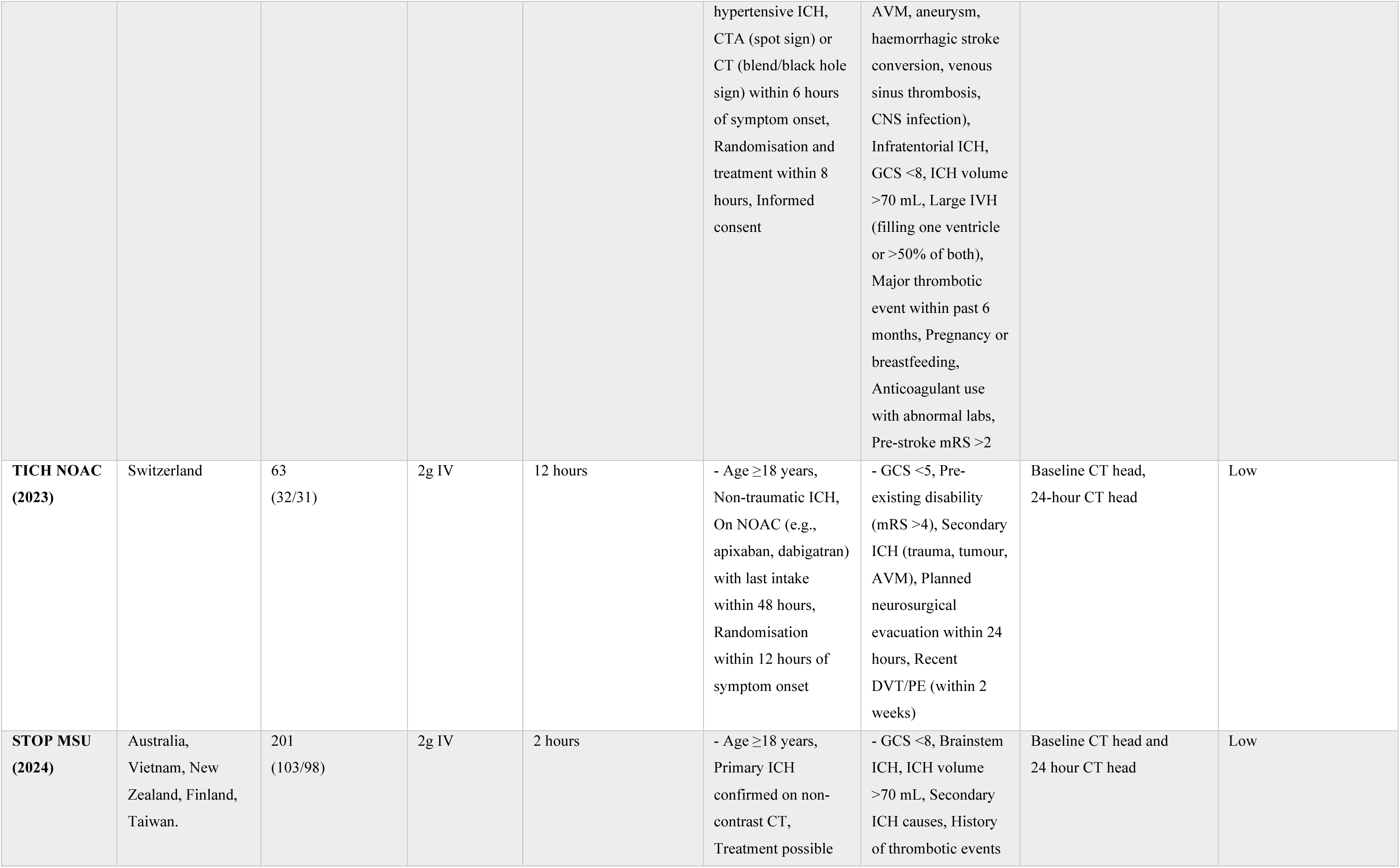
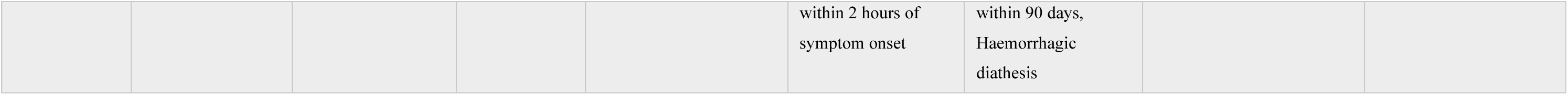
Characteristics of included randomised controlled trials. Summary of the five trials contributing individual participant data to the meta-analysis. Trials are described by country, sample size, tranexamic acid dosing regimen, treatment time window, imaging protocol, and risk of bias assessment.

| Study (Year) | Country | Sample Size<br>(tranexamic acid /<br>Control) | Tranexamic<br>acid Dose &<br>Route | Onset to Treatment<br>(hrs) | Key Inclusion<br>Criteria | Key Exclusion<br>Criteria | Imaging Criteria | Risk of Bias |
| --- | --- | --- | --- | --- | --- | --- | --- | --- |
| **TICH 2**<br>**(2017)** | UK, Denmark,<br>Georgia, Hungary,<br>Ireland, Italy,<br>Malaysia, Poland,<br>Spain, Sweden,<br>Switzerland,<br>Turkey. | 2325<br>(1161/1164) | 2g IV | 8 hours | - Age $\geq 18$ years,<br>Spontaneous ICH<br>confirmed on CT,<br>Randomised within 8<br>hours of symptom<br>onset | - Secondary ICH<br>(trauma, tumour,<br>AVM), Planned<br>surgical evacuation,<br>GCS $< 5$, Known<br>thromboembolic<br>event within previous<br>90 days,<br>Contraindication to<br>tranexamic acid | Baseline CT head +/- CTA,<br>24-hour CT head | Low |
| **STOP AUST**<br>**(2020)** | Australia, Finland,<br>Taiwan | 100<br>(50/50) | 2g IV | 4.5 hours | - Age $\geq 18$ years, ICH<br>on CT with spot sign<br>on CTA,<br>Randomisation<br>within 4.5 hours of<br>onset, GCS $> 7$, ICH<br>volume $< 70$ mL | -GCS $< 5$, Pre-existing disability<br>(mRS $> 4$), Secondary<br>ICH (trauma, tumour,<br>AVM), Planned<br>neurosurgical<br>evacuation within 24<br>hours, Recent<br>DVT/PE (within 2<br>weeks) | Baseline CT head and CTA<br>scan, 24-hour CT head | Low |
| **TRAIGE**<br>**(2021)** | China | 171<br>(89/82) | 2g IV | 8 hours | - Age 18-79 years,<br>spontaneous | - Secondary ICH<br>(trauma, tumour, | Baseline CT head +/- CTA,<br>24-hour CT head | Low |
|  |  |  |  |  | hypertensive ICH, CTA (spot sign) or CT (blend/black hole sign) within 6 hours of symptom onset, Randomisation and treatment within 8 hours, Informed consent | AVM, aneurysm, haemorrhagic stroke conversion, venous sinus thrombosis, CNS infection), Infratentorial ICH, GCS <8, ICH volume >70 mL, Large IVH (filling one ventricle or >50% of both), Major thrombotic event within past 6 months, Pregnancy or breastfeeding, Anticoagulant use with abnormal labs, Pre-stroke mRS >2 |  |  |
| **TICH NOAC (2023)** | Switzerland | 63 (32/31) | 2g IV | 12 hours | - Age ≥18 years, Non-traumatic ICH, On NOAC (e.g., apixaban, dabigatran) with last intake within 48 hours, Randomisation within 12 hours of symptom onset | - GCS <5, Pre-existing disability (mRS >4), Secondary ICH (trauma, tumour, AVM), Planned neurosurgical evacuation within 24 hours, Recent DVT/PE (within 2 weeks) | Baseline CT head, 24-hour CT head | Low |
| **STOP MSU (2024)** | Australia, Vietnam, New Zealand, Finland, Taiwan. | 201 (103/98) | 2g IV | 2 hours | - Age ≥18 years, Primary ICH confirmed on non-contrast CT, Treatment possible within 2 hours of<br>symptom onset | - GCS <8, Brainstem ICH, ICH volume >70 mL, Secondary ICH causes, History of thrombotic events within 90 days,<br>Haemorrhagic<br>diathesis | Baseline CT head and 24 hour CT head | Low |

The two-stage aggregate data analysis incorporated data from Arumugam et al 2014 (n=30) and Ni et al 2020 (n=150), extending the evidence base^10,17^. The TANICH trial (n=60) was identified, but its published outcome data was not suitable for inclusion^11^. The ITCH trial (n=96) were unable to share their data with us as they were awaiting publication of their primary results (<u>NCT04742205</u>).

### Baseline Characteristics

A total of 2,860 patients were included in the analysis, with 1,435 assigned to tranexamic acid arm and 1425 assigned to placebo arm. Baseline demographic and clinical characteristics were similar between the tranexamic acid and placebo groups and are summarised in Table 2. The mean age was 68 (14.1) years in both groups, and the proportion of female patients was 760/1435 (54.1%) and 770/1425 (53.0%), respectively. The median premorbid modified Rankin Scale score was 0 (0-1) in both arms. Common vascular risk factors including hypertension, diabetes, ischaemic heart disease, prior stroke or TIA, and statin or antiplatelet use were similarly distributed across groups.

**Table 2.** Participant-level baseline demographic, clinical and radiographic characteristics by treatment group.

| Baseline Patient Characteristics Variables | N | Tranexamic acid<br>(n=1,435) | Placebo<br>(n=1,425) |
| --- | --- | --- | --- |
| Age, years; mean (SD)* | 2860 | 68.2 (14.1) | 68.0 (14.1) |
| Female Sex (n, %) | 2860 | 760 (54) | 770 (53) |
| Premorbid mRS; Median (IQR)* | 2860 | 0 (0-1) | 0 (0-1) |
| Medical History (n, %) |  |  |  |
| Hypertension | 2689 | 830 (61.7) | 842 (62.7) |
| Diabetes | 2858 | 213 (14.9) | 185 (13.0) |
| Ischemic Heart Disease | 2796 | 127 (9.1) | 112 (8.1) |
| Prior Stroke or Transient Ischemic Attack | 2856 | 199 (13.9) | 181 (12.7) |
| Antiplatelet Use | 2857 | 360 (25.2) | 330 (23.2) |
| Statin use | 2346 | 359 (25.2) | 354 (24.9) |
| NIHSS; Median (IQR)* | 2839 | 13 (7-18) | 12 (6-18) |
| Glasgow Coma Scale; Median (IQR) | 2840 | 15 (12-15) | 15 (12-15) |
| Systolic Blood Pressure; median (IQR)* | 2856 | 170 (153 – 189) | 171 (153-190) |
| Diastolic Blood Pressure; median (IQR) | 2855 | 91.5 (80-105) | 92 (81-105) |
| Weight; median (IQR) | 1997 | 73.4 (63.5 – 85.0) | 75 (65-88.7) |
| Glucose; median (IQR) | 2357 | 6.7 (5.7-8.1) | 6.5 (5.7-7.8) |
| Onset to randomisation; median (IQR)* | 2857 | 3.5 (2.4 – 5.0) | 3.5 (2.3-4.9) |
| Onset to treatment; median (IQR) | 2835 | 3.8 (2.8-5.5) | 3.8 (2.7 – 5.4) |
| ICH cause (n, %) | 2858 |  |  |
| - Cerebral Amyloid Angiopathy |  | 63 (4.4) | 63 (4.4) |
| - Hypertension |  | 755 (52.6) | 768 (54) |
| - Secondary causes |  | 45 (3.1) | 57 (4) |
| - Unknown |  | 572 (39.9) | 535 (37.6) |
| Baseline Radiological Variables |  |  |  |
| Onset to CT scan; median (IQR) | 2860 | 2.17 (1.5 – 2.2) | 2.15 (1.4 – 3.6) |
| Baseline Intraparenchymal Volume, mL; median (IQR)* | 2807 | 14.1 (6.0-31.9) | 12.8 (5.3-30.5) |
| Intraventricular Haemorrhage Presence at Baseline (n, %) | 2854 | 412 (28.7) | 404 (28.4) |
| Baseline Intraventricular Volume, mL; median (IQR) | 1227 | 2.1(0-9.7) | 1.7 (0-8.7) |
| Intracerebral Haemorrhage Location(n, %)* | 2829 |  |  |
| Supratentorial Deep |  | 933 (65.9) | 929 (65.9) |
| Supratentorial Lobar |  | 406 (28.8) | 410 (28.9) |
| Infratentorial |  | 76 (5.4) | 74 (5.2) |
| **CT angiography spot sign (n, %)** | 739 | 142 (38.2) | 138 (37.6) |
| **CT non contrast Blackhole sign (n, %)** | 2331 | 205 (17.4) | 200 (17.3) |
| **CT island sign (n, %)** | 1386 | 111 (15.6) | 111 (16.5) |
| **CT Blend sign (n, %)** | 2645 | 263 (19.8) | 253 (19.2) |

Data are n (%), mean (SD), or median (IQR). NIHSS=National Institutes of Health Stroke Scale. \*Minimisation criteria.

Clinical severity on admission, including NIHSS score and GCS, did not differ significantly between groups. The median NIHSS score was 13 (7-18) in the tranexamic acid group and 12 (6-18) in the placebo group, while the median GCS was 15 (12-15) in both groups. Blood glucose, weight, systolic and diastolic blood pressure readings were also comparable. Onset to randomisation was similar between groups, with a median of 3.5 (2.4-5.0) hours in the tranexamic acid group and 3.5 (2.3-4.9) hours in the placebo group.

Radiological characteristics were likewise similar. Median onset to CT scan was similar between groups, with a median of 2.17 (1.5-2.2) hours in the tranexamic acid group and 2.14 (1.4-3.6) in placebo group. The median baseline haematoma volume was 14.1 (6.0-31.9) mL in the tranexamic acid group and 12.8 (5.3-30.5) mL in the placebo group. Intraventricular haemorrhage was present at baseline in approximately 28% of patients in both groups. Haematoma location was distributed similarly across supratentorial deep, supratentorial lobar, and infratentorial regions.

Imaging markers predictive of haematoma expansion, including CT angiography spot sign, non-contrast dot sign, island sign, and blend sign, were also evenly distributed.

### Primary and Secondary Outcomes

The primary outcome was functional status at 90 days, measured using the modified Rankin Scale. 757/1423 (53.2%) patients assigned to tranexamic acid had a worse functional outcome compared with 759/1415 (53.6%) assigned to placebo. The adjusted common odds ratio (adjusted common OR) in the ordinal analysis testing for a shift towards worse modified Rankin Score was 0.93 (95% CI 0.81 to 1.07; p = 0.30) (Table 3). The proportional odds assumption was held (p = 0.08). The likelihood ratio test comparing fixed and random-effects models showed no evidence of meaningful between-trial heterogeneity (χ² = 42102.96, p < 0.001), supporting consistency of treatment effect across trials. The modified Rankin Score shift plot confirmed the similarity in functional outcome distribution, with no significant directional effect (Supplementary Figure 2). Summary of findings assessed on GRADE suggests that tranexamic acid probably does not reduce long-term dependence (moderate certainty) (Supplementary Table 1).

**Table 3.**
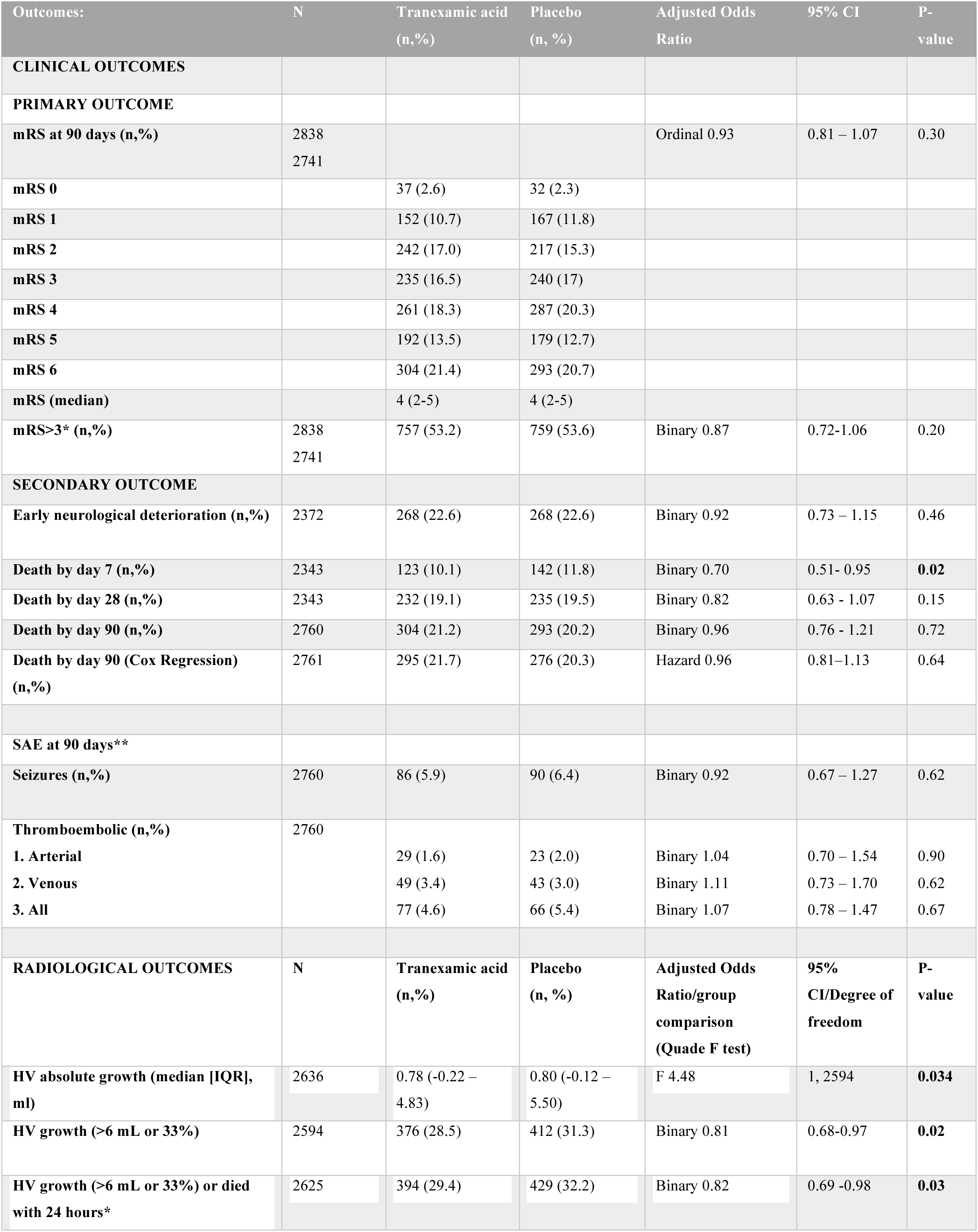
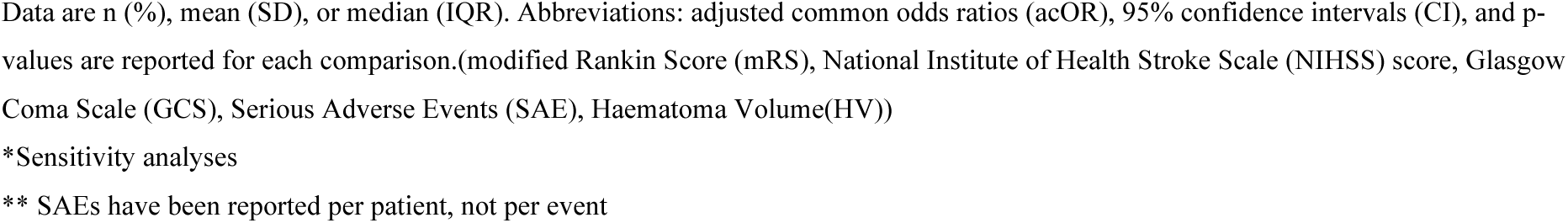
One stage individual patient data analysis. Primary, secondary, and radiological outcome measures by treatment group.

| Outcomes: | N | Tranexamic acid<br>(n,%) | Placebo<br>(n, %) | Adjusted Odds<br>Ratio | 95% CI | P-value |
| --- | --- | --- | --- | --- | --- | --- |
| **CLINICAL OUTCOMES** |  |  |  |  |  |  |
| **PRIMARY OUTCOME** |  |  |  |  |  |  |
| mRS at 90 days (n,%) | 2838<br>2741 |  |  | Ordinal 0.93 | 0.81 – 1.07 | 0.30 |
| mRS 0 |  | 37 (2.6) | 32 (2.3) |  |  |  |
| mRS 1 |  | 152 (10.7) | 167 (11.8) |  |  |  |
| mRS 2 |  | 242 (17.0) | 217 (15.3) |  |  |  |
| mRS 3 |  | 235 (16.5) | 240 (17) |  |  |  |
| mRS 4 |  | 261 (18.3) | 287 (20.3) |  |  |  |
| mRS 5 |  | 192 (13.5) | 179 (12.7) |  |  |  |
| mRS 6 |  | 304 (21.4) | 293 (20.7) |  |  |  |
| mRS (median) |  | 4 (2-5) | 4 (2-5) |  |  |  |
| mRS>3* (n,%) | 2838<br>2741 | 757 (53.2) | 759 (53.6) | Binary 0.87 | 0.72-1.06 | 0.20 |
| **SECONDARY OUTCOME** |  |  |  |  |  |  |
| Early neurological deterioration (n,%) | 2372 | 268 (22.6) | 268 (22.6) | Binary 0.92 | 0.73 – 1.15 | 0.46 |
| Death by day 7 (n,%) | 2343 | 123 (10.1) | 142 (11.8) | Binary 0.70 | 0.51- 0.95 | **0.02** |
| Death by day 28 (n,%) | 2343 | 232 (19.1) | 235 (19.5) | Binary 0.82 | 0.63 - 1.07 | 0.15 |
| Death by day 90 (n,%) | 2760 | 304 (21.2) | 293 (20.2) | Binary 0.96 | 0.76 - 1.21 | 0.72 |
| Death by day 90 (Cox Regression)<br>(n,%) | 2761 | 295 (21.7) | 276 (20.3) | Hazard 0.96 | 0.81–1.13 | 0.64 |
| SAE at 90 days** |  |  |  |  |  |  |
| Seizures (n,%) | 2760 | 86 (5.9) | 90 (6.4) | Binary 0.92 | 0.67 – 1.27 | 0.62 |
| Thromboembolic (n,%) | 2760 |  |  |  |  |  |
| 1. Arterial |  | 29 (1.6) | 23 (2.0) | Binary 1.04 | 0.70 – 1.54 | 0.90 |
| 2. Venous |  | 49 (3.4) | 43 (3.0) | Binary 1.11 | 0.73 – 1.70 | 0.62 |
| 3. All |  | 77 (4.6) | 66 (5.4) | Binary 1.07 | 0.78 – 1.47 | 0.67 |
| **RADIOLOGICAL OUTCOMES** | **N** | **Tranexamic acid**<br>**(n,%)** | **Placebo**<br>**(n, %)** | **Adjusted Odds**<br>**Ratio/group**<br>**comparison**<br>**(Quade F test)** | **95%**<br>**CI/Degree of**<br>**freedom** | **P-**<br>**value** |
| HV absolute growth (median [IQR],<br>ml) | 2636 | 0.78 (-0.22 –<br>4.83) | 0.80 (-0.12 –<br>5.50) | F 4.48 | 1, 2594 | **0.034** |
| HV growth (>6 mL or 33%) | 2594 | 376 (28.5) | 412 (31.3) | Binary 0.81 | 0.68-0.97 | **0.02** |
| HV growth (>6 mL or 33%) or died<br>with 24 hours* | 2625 | 394 (29.4) | 429 (32.2) | Binary 0.82 | 0.69 -0.98 | **0.03** |

Data are n (%), mean (SD), or median (IQR). Abbreviations: adjusted common odds ratios (acOR), 95% confidence intervals (CI), and p-values are reported for each comparison.(modified Rankin Score (mRS), National Institute of Health Stroke Scale (NIHSS) score, Glasgow Coma Scale (GCS), Serious Adverse Events (SAE), Haematoma Volume(HV))
\*Sensitivity analyses
\*\* SAEs have been reported per patient, not per event

Early neurological deterioration, defined as a worsening in either NIHSS increase by 4 or GCS drop by 2 within 24 hours, occurred in 268/1188 (22.6%) of patients receiving tranexamic acid and 268/1184 (22.6%) of those receiving placebo, yielding an adjusted OR of 0.92 (95% CI 0.73 to 1.15; p = 0.46).

All-cause mortality at day 7 was significantly lower in the tranexamic acid group compared to placebo 123/1213 (10.1%) vs. 142/1203 (11.8%); adjusted OR 0.70, 95% CI 0.51 - 0.95; p = 0.03. However, this difference was not sustained at day 28 232/1213 (19.1%) vs. 235/1203 (19.5%); adjusted OR 0.82, 95% CI 0.63 to 1.07; p = 0.15) or at day 90 304/1435 (21.0%) vs. 293/1425 (20.2%); adjusted OR 0.96, 95% CI 0.76 to 1.21; p = 0.72, high certainty). The cox regression survival analysis showed that cumulative survival was similar between the tranexamic acid and placebo groups over the 90-day follow-up period (Supplementary Figure 4). The tranexamic acid group shows a slightly higher survival probability at most time points, but the difference is minimal. The hazard ratio(HR) for death with tranexamic acid compared to placebo is insignificant (adjusted HR 0.96 (95% CI 0.81–1.13; p = 0.64)).

There was a statistically significant difference in haematoma volume absolute growth difference between treatment groups after adjusting for baseline covariates using the Quade non-parametric ANCOVA (F(1, 2594) = 4.48, p = 0.03). This significant difference was consistent across categorical definitions of haematoma expansion: HV growth >6 ml or >33% from baseline occurred in 376/1321 (28.5%) in the tranexamic acid group versus 412/1315 (31.3%) in the placebo group (adjusted OR 0.81, 95% CI 0.68–0.97; p = 0.02, high certainty).

Serious adverse events at 90 days were comparable between the tranexamic acid and placebo groups. Seizures occurred in 86/1435 (5.9%) of patients receiving tranexamic acid and 90/1425 (6.4%) receiving placebo (adjusted OR 0.92, 95% CI 0.67–1.27; p = 0.62, high certainty). Thromboembolic events occurred in 77/1435 (4.6%) of the tranexamic acid and 66/1425 (5.4%) of the placebo group (adjusted OR 1.06, 95% CI 0.77–1.45; p = 0.72, high certainty). When disaggregated, arterial events were reported in 29/1435 (2%) and 23/1425 (1.6%), and venous events in 49/1435 (3.4%) and 43/1425 (3.0%) of patients in the tranexamic acid and placebo groups, respectively. None of these differences were statistically significant. These results suggest that tranexamic acid was not associated with an increased risk of seizures or thromboembolic complications at 90 days.

In the study level aggregate data analysis the proportion of patients with an shift towards worse functional outcome using the modified Rankin Score at 90 days showed no significant difference between the tranexamic acid and placebo groups (OR 0.90, 95% CI 0.78 to 1.03; I² = 0%, 5 trials, 2838/3040 (93%)).For mortality, there was no significant difference in death at 90 days (OR 1.04, 95% CI 0.87 to 1.25; I² = 0%, 7 trials, 3040/3040 (100%)). Tranexamic acid was associated with a statistically significant reduction in haematoma expansion compared to placebo (OR 0.84, 95% CI 0.72 to 0.99; I² = 0%, 7 trials, 3040/3040 (100%)). With regards to safety outcomes, there was no significant difference in the incidence of thromboembolic adverse events (OR 1.16, 95% CI 0.83 to 1.63; I² = 0%, 6 trials, 3010/3040 (99%))(Figure 2). There was no significant heterogeneity observed across any of the outcomes, in-keeping with the primary analysis (Figure 2).

**Figure 2.**
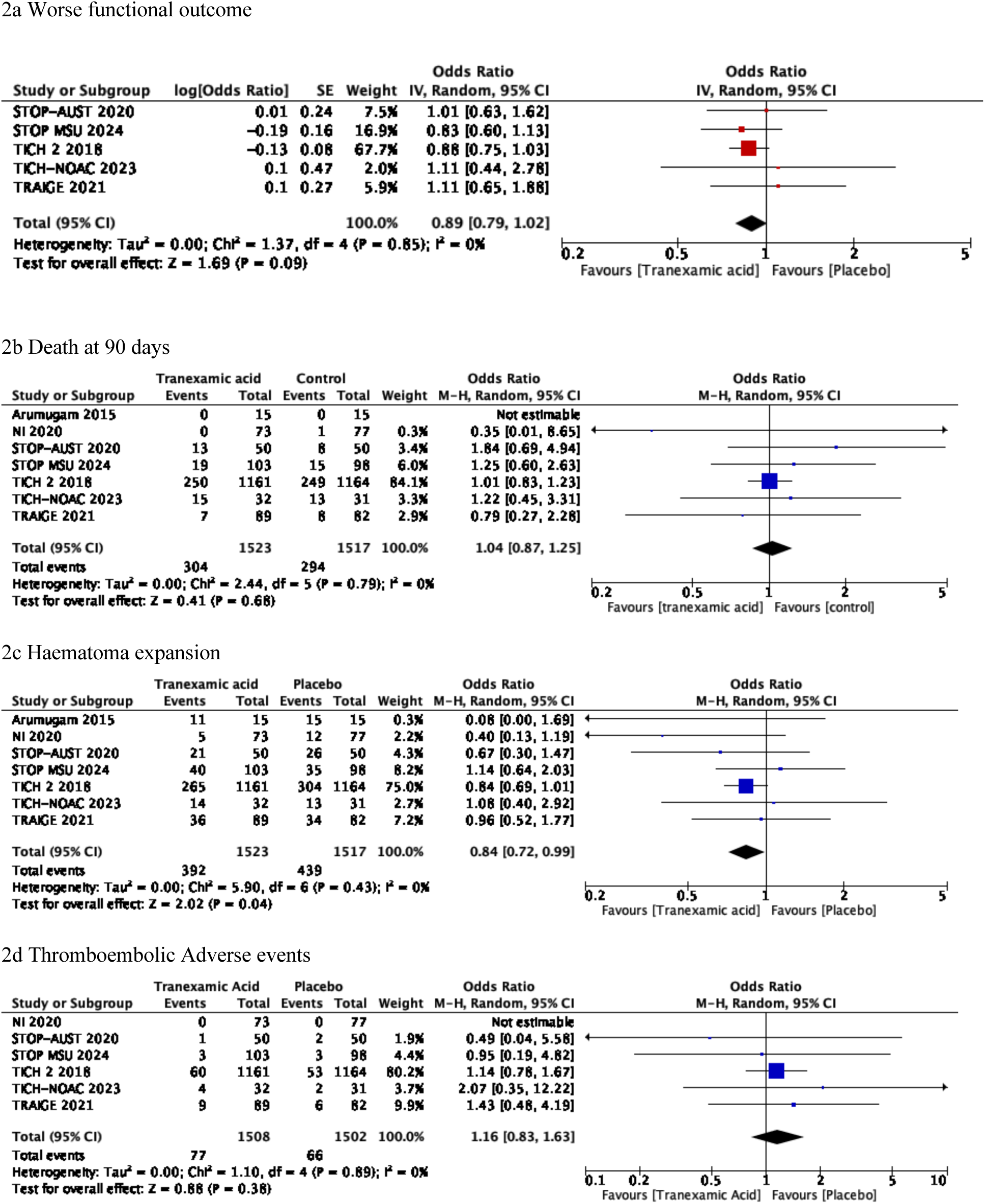
Aggregate study-level meta-analyses.

### Subgroup Analyses

Prespecified subgroup analyses were performed to assess whether the effect of tranexamic acid varied across clinical and radiological strata. No statistically significant interaction was observed in any subgroup, including by age, sex, blood pressure, NIHSS score, onset-to-treatment time, haematoma volume or location, presence of intraventricular haemorrhage, prior antiplatelet or anticoagulant use, or CT/CTA features such as spot sign and non-contrast haematoma markers. Any possible signals noted were not supported by an interaction effect across the variable and may represent a chance finding (Figure 3).

**Figure 3.**
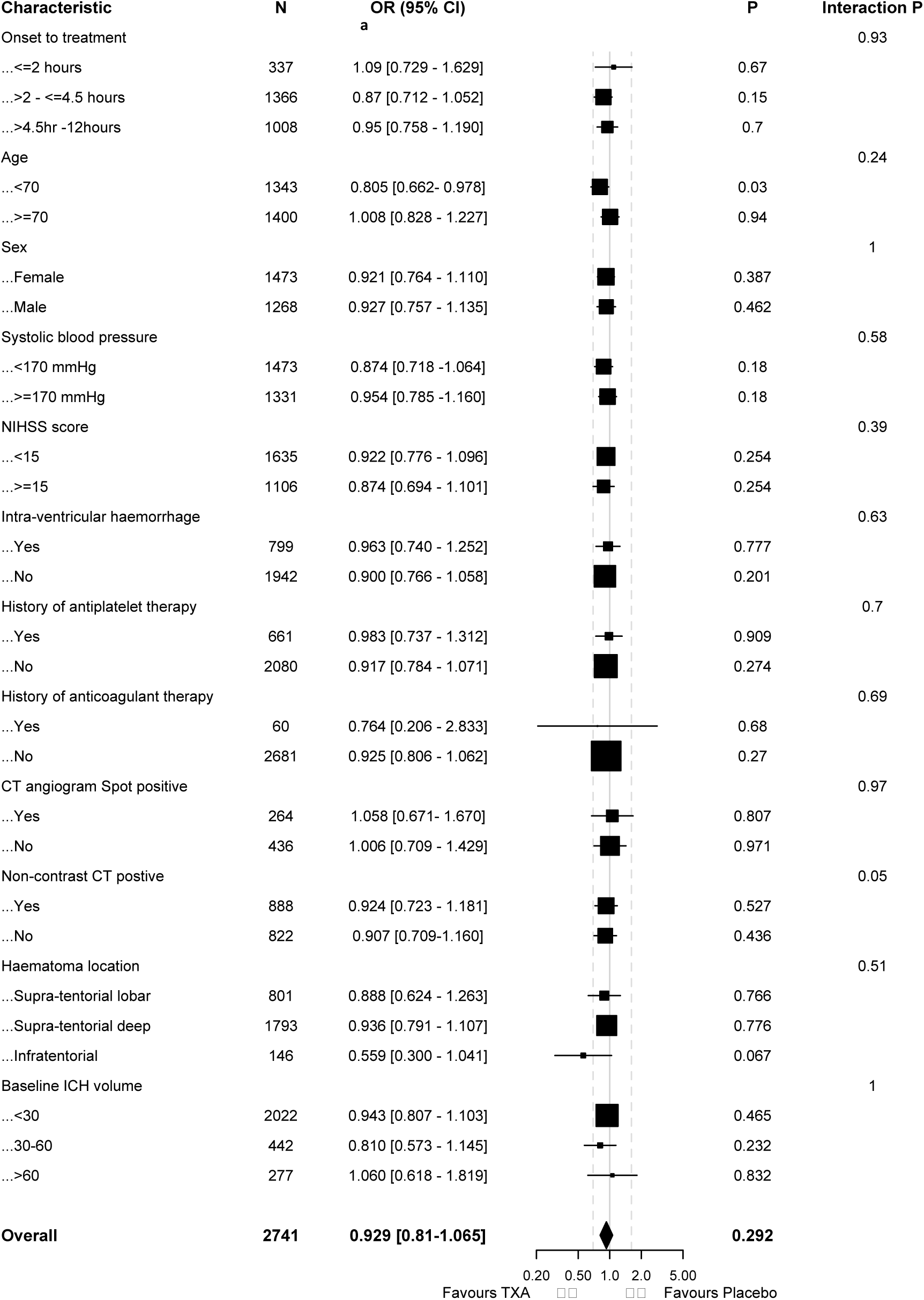
One-stage Subgroup analysis of the effect of tranexamic acid on mRS at 90 days. Forest plot of adjusted common odds ratios (with 95% confidence intervals) for the primary outcome across prespecified subgroups.

### Sensitivity Analyses

There were no single-centre trials, trials with high risk of bias, or trials lacking a published protocol included in the one stage individual patient data meta-analysis.

Binary analysis of the modified Rankin Score, categorising outcomes into favourable (mRS ≤3) and unfavourable (modified Rankin Score >3), showed no significant treatment effect. Poor outcome occurred in 757/1423 (53.2%) of patients in the tranexamic acid group compared with 759/1415 (53.6%) in the placebo group (adjusted OR 0.87, 95% CI 0.71–1.06; p = 0.20).

Sensitivity analysis for haematoma expansion accounted for participants who died within the first 24 hours, as these patients would not have undergone repeat imaging. When including early death (within 24 hours) in the composite outcome, the event occurred in 394/1338 (29.4%) in the tranexamic acid group compared to 429/1331 patients (32.2%) in the placebo group (adjusted OR 0.82, 95% CI 0.69–0.98; p = 0.03), again showing a statistically significant reduction with tranexamic acid.

A fixed-effect model was performed as a sensitivity analysis due to the lack of formal statistical evidence for between-trial heterogeneity in baseline outcome risk. Results were consistent with the primary analysis. The adjusted OR for modified Rankin Score shift at 90 days was 1.11 (95% CI 0.97–1.28; p = 0.13), and for poor outcome (modified Rankin Score >3) was 0.87 (95% CI 0.71–1.09; p = 0.20). The reduction in day 7 mortality persisted (adjusted OR 0.72, 95% CI 0.55–0.95), though remained non-significant at later time points. Haematoma expansion results were also consistent (adjusted OR 0.82, 95% CI 0.69–0.97; p = 0.03). Adverse events rates were also no different between tranexamic acid or placebo groups (Supplementary Table 2).

## Discussion

In this individual participant data meta-analysis of five RCTs investigating tranexamic acid in spontaneous ICH, we found no significant difference in functional outcome at 90 days. The distribution of modified Rankin Score scores and the proportion of patients with poor outcomes were similar between the tranexamic acid and placebo groups. Tranexamic acid was associated with a reduced haematoma expansion and early mortality at day 7; however, this mortality benefit was not sustained at 28 or 90 days. These findings were consistent across clinical and radiological subgroups. As well as robust in sensitivity analyses using fixed- and random-effects models.

These findings suggest that while tranexamic acid reduces haematoma expansion, the effect may not be sufficient to improve long-term outcomes in unselected patients. The transient survival benefit observed at day 7 could reflect early stabilisation of the haematoma, possibly through prevention of rebleeding or further expansion^18,19^. However, the absence of sustained benefit is less clear. Potential explanations include inadequate sample size, treatment being given too late during injury, irreversible baseline damage, or the modest clinical impact of reductions in haematoma volume. Longer-term outcomes are also likely to be driven by other factors such as perihaematomal oedema, haematoma location, and intraventricular extension ^20–22^.

Several systematic reviews and meta-analyses have previously assessed the efficacy of tranexamic acid in spontaneous ICH. Most of these reviews, including a Cochrane review published in 2023, relied on aggregate-level data and reported that while tranexamic acid appeared to reduce haematoma expansion, there was no convincing evidence of improved functional outcomes or mortality benefits ^23–29^. Radiological markers such as the CT angiogram spot sign and NCCT markers did not show significance in any subgroup in our individual patient data meta-analysis however in previous studies spot sign positive CTA has favoured tranexamic acid in with treatment <4.5 hours and a post-hoc analysis in TICH 2 showed positive markers in NCCT had better mortality and morbidity outcomes at 90 days^30,31^.

While previous reviews reported no effect of tranexamic acid on functional outcome, this analysis adds precision and confirms modest early benefits ^23–27,29^. Although the current evidence suggests that tranexamic acid alone is unlikely to shift long-term outcomes, in the context of care bundles such as those implemented in INTERACT3 or ABC-ICH^32,33^, tranexamic acid may provide additive benefit when delivered alongside blood pressure control, minimally invasive neurosurgical intervention^34–36^, and anticoagulation reversal. Tranexamic acid’s low cost, good safety profile, and ease of administration strengthen its case for inclusion in such protocols if ongoing trials such as INTRINSIC ^37^ and TICH-3 (ISRCTN97695350) confirm clinical benefit in ICH treatment.

The main strength of this study is the use of individual participant data across five RCTs (90% of all available individual patient data) provided sufficient power to detect modest effects and allowed for harmonisation of outcome definitions, covariate adjustments, and subgroup analysis.

However, some limitations must be acknowledged including dominance of the TICH-2 trial in the dataset, missing data was not imputed, and lack of individual patient data from some smaller studies. Although site-level clustering was not accounted for, trial-level random effects showed no evidence of heterogeneity, supporting the consistency of treatment effect across included trials. The possibility of selection bias or limited generalisability should also be acknowledged.

In summary, tranexamic acid is safe and reduces early haematoma growth and short-term mortality, but without improving long-term outcomes. Larger ongoing trials such as TICH-3, and INTRINSIC will be important in clarifying tranexamic acid’s role, particularly in patients treated in less than 4.5 hours, on anticoagulants, and with expansion-prone bleeds^37,38^. Even small treatment effect may have public health significance, given the global burden of ICH.

## Data Availability

Individual participant data and the data dictionary will not be made available. Requests should be sent to.

## Acknowledgements

We would like to thank all our international collaborators for contributing and supporting this individual meta-analysis.

## Conflicts of Interest

CSM: Involved in the ongoing TICH-3 trial.

ZKL: Contributor to the TICH-2 trial.

LW: Contributor to TICH-2 and TICH-3 trials.

JL: Involved in the TRIAGE trial.

LL: Holds research grant and Involved in the TRIAGE trial.

NY: Holds research grants and involved in STOP-AUST and STOP-MSU trials. Received honoraria for educational activities from Eli Lilly and Novo Nordisk.

CM, HZ, SD, GD, AM: Involved in STOP-AUST and STOP-MSU trials. AM: Receives consulting fees from Boehringer-Ingelheim and sits on the committee board for World Stroke Forum.

JP: Involved in the INTRINSIC trial; President of the World Stroke Congress.

RD: Contributor to TICH-2 and TICH-3 trials.

DS and AP: Holds research and involved in TICH NOAC

PMB: Contributor to TICH-2 and TICH-3 trials. Holds NIHR and BHF grants paid to the University of Nottingham. Receives lecture/consulting fees from Phagenesis, Roche, DiaMedica; holds stock options in CoMind.

Industry Committee Co-Chair, WSC.

RA-SS: Involved in grant applications and delivery of TICH-2 and TICH-3 trials. Holds two NIHR grants paid to the University of Edinburgh. Past President, British & Irish Association of Stroke Physicians.

NS: Holds research grants for TICH-2, DASH, and TICH-3 trials.

## Contributions

Chaamanti Menon: protocol development, statistical analysis, data interpretation, manuscript writing, risk of bias and GRADE analysis.

Zhe Kang Law: protocol development.

Lisa Woodhouse: statistical analysis and data interpretation.

Jingyi Liu: risk of bias and GRADE analysis.

Chloe Mutimer: collaborator.

Michael Desborough: protocol development, risk of bias analysis.

Liping Liu: collaborator.

Alexandros Polymeris: collaborator.

Atte Meretoja: collaborator.

Nawaf Yassi: collaborator.

Henry Zhao: collaborator.

Stephen Davis: collaborator.

Geoffrey Donnan: collaborator.

Jeyaraj Pandian: collaborator.

Rob A. Dineen: collaborator.

David Seiffge: collaborator.

Rustam Al-Shahi Salman: protocol development.

Philip M Bath: protocol development, statistical analysis, data interpretation, manuscript writing.

Nikola Sprigg: protocol development, statistical analysis, data interpretation, manuscript writing.

All authors had full access to all the data in the study and had final responsibility for the decision to submit for publication.

## Non-standard Abbreviations and Acronyms

ICH: Intracerebral Haemorrhage
GCS: Glasgow Coma Scale
NIHSS: National Institute of Health Stroke Scale
WHO ICTRP: WHO International Clinical Trials Registry Platform

